# Prognostic Model Construction Based on Platinum-Free Interval in Ovarian Cancer and Its Implication for Chemotherapy Resistance

**DOI:** 10.1101/2024.12.02.24318249

**Authors:** Yang Zhang, Jihui Kang, Chuling Wu, Huishan Xu, Qin Ling, Hao Tan, Zuwei Zhang, Weipeng He, Shuzhong Yao, Langyu Gu, Guofen Yang

## Abstract

**Background:** While platinum sensitivity and resistance have long been central to treatment decisions in high-grade serous ovarian cancer (HGSOC), these categories are increasingly questioned in real-world clinical settings. This study seeks to develop a prognostic model based on platinum-free interval (PFI) as a reliable indicator of patient prognosis, with additional exploration of chemotherapy resistance-related genes and pathways.

**Methods:** 70 HGSOC patients with varied gene expression profiles and corresponding clinical information of platinum-based chemotherapy responses were analysed. We first identified PFI-related genes (PRGs) that constituted a predictive signature for HGSOC by using univariate COX and LASSO regression analysis. We determined the optimal PFI indicative using linear correlation equations between gene expression levels and PFI. This time point was then employed to categorize patients into cohorts with good and poor prognosis, followed by an analysis of differentially expressed genes (DEGs) and their enriched pathways. Additionally, we utilized public available drug database to evaluate chemotherapeutic agents effective against the poor prognosis group.

**Results:** A signature comprising 10 PRGs (TUBA4A, ENSG00000232325.3, ENSG00000268080.1, KCNK9, ENSG00000230567.3, CST6, KNTC1, LINC02167, ENSG00000267469.1, NKAIN4) was established. Patients within the high-risk category defined by this signature exhibited a poorer prognosis and earlier recurrence than low-risk group. The prognostic model had a robust accuracy in predicting prognosis with an area under curve value >0.90. We estimated a PFI threshold of 22.37 months, which serves as a cutoff point to further differentiate groups with good and poor prognosis. KEGG pathways enrichment analysis revealed that taurine and hypotaurine metabolism, melanogenesis, Cushing syndrome, and mTOR signaling pathways were enriched in the poor prognosis group. We also performed drug resistance assessment and found that patients from the poor prognosis group were more sensitive to anti-cancer drugs such as Pevonedistat and GDC0810 than the good prognosis group.

**Conclusions:** Our study constructed a prognostic model based on PFI for HGSOC and further explored its implications for chemotherapy resistance. These findings could enhance clinical applications and inform novel anticancer therapeutic strategies targeting HGSOC.

## Background

High-grade serous ovarian cancer (HGSOC) represents one of gynecologic oncology’s most lethal malignancies(1,2). With advancements in high-throughput genetic testing, the management strategies for HGSOC have undergone significant progress(3). Yet, the cornerstone of treating this cancer is still surgical intervention alongside platinum-based chemotherapy(4). Nevertheless, cancer persistently recurs and become less responsive to platinum drugs inevitably. The challenge posed by developing solutions against platinum resistance remains an insurmountable obstacle(5). The assessment of platinum-free interval (PFI) stands out as a crucial functional predictive indicator guiding therapeutic decisions for chemotherapy responses in HGSOC(6). Currently, the NCCN guidelines define patients with PFI less than 6 months as platinum-resistant, and patients with non-remission after two courses of platinum-based chemotherapy in the initial treatment as platinum-refractory (in this paper, both are called platinum resistance). And patients classified under ‘platinum-sensitive’ are whose disease recur more than 6 months. This platinum responses time threshold classification is important for clinical treatment, as ‘platinum-sensitive’ patients retain access to platinum-based chemotherapeutic regimens, but ‘platinum-resistant’ patients face restricted range of poorly effective non-platinum therapies (7).

However, ongoing clinical studies keep challenging the current PFI-based criteria for distinguishing platinum-resistant from platinum-sensitive. For example, retrospective analyses like SOLO2 and PAOLA1 trials suggest that incorporating maintenance therapies such as bevacizumab and PARP inhibitors could prolong PFI to a certain extent, leading to a subset of platinum-resistant patients being incorrectly defined as platinum-sensitive and consequently receiving platinum-based chemotherapy that they should not administered under current guidelines (8,9). On the other hand, an Australian Ovarian Cancer Study retrospective analysis revealed that even for patients who were classified as platinum-resistant (PFI < 6 months) that should not continue using platinum-based therapy, can still benefit from platinum-based therapy with enhanced overall survival (10). The NCCN guidelines (2024.v1) also begin to emphasize that current definitions pertaining to ‘platinum-sensitive’/’platinum-resistant’ lack precision (11). Therefore, although researchers increasingly recognize that the current PFI threshold for distinguishing platinum-resistant from platinum-sensitive is inadequate, there is still no better solution available yet(12).

In fact, platinum resistance is a complex biological phenomenon with its molecular genetic basis(13). Many existing studies have revealed that changes in the expression levels of genes can play important roles in the response of tumour cells to platinum chemotherapy drugs(14–16). Additionally, in our recent study using single-cell transcriptomics to investigate key cell populations and their core regulatory networks for chemotherapy resistance in HGSOC, we discovered that the core co-expression regulatory network related to chemotherapy resistance in cancer cells is actually present in resistant individuals before chemotherapy treatment(17). These suggest that gene expression profile changes in pre-chemotherapy samples may affect chemotherapy responses and help predict prognosis for HGSOC patients. These research foundations lay the groundwork for exploring the correlation between gene expression levels in pre-chemotherapy samples and prognosis based on PFI. The corresponding differentially expressed genes (DEGs) are likely to be closely related to chemotherapy response and may therefore serve as potential therapeutic targets or prognostic biomarkers.

## Methods

### 1. Data acquisition

The transcriptome data and expression matrix for HGSOC patients were obtained from the Gene Expression Omnibus (GEO) database (GSE102073) (18). After excluding patients without PFI or gene expression information, a total of 70 HGSOC patients were included for the following analyses.

### 2. Establishment and verification of the PFI-related genes (PRGs) prognostic signature

PFI is defined as the duration between the last administration of platinum-containing chemotherapy and the occurrence of disease progression. Disease progression is determined using RECIST (Response Evaluation Criteria in Solid Tumor) criteria, which includes the longest tumor diameter and an increase of ≥20%, the emergence of new lesions, or the unmeasurable progression of malignant lesions(19).

The "survival" R package was utilized to identify genes associated with PFI, with a significance level of *p* < 0.05 determined by univariate Cox regression analysis. Subsequently, least absolute shrinkage and selection operator (LASSO) regression analysis was performed to filter genes using 100-fold cross-validation. The risk score was calculated using the formula:

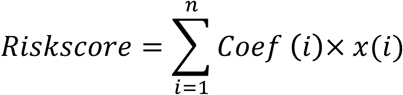

where x(i) and Coef (i) represent the expression levels of each gene and their respective regression coefficient. HGSOC patients were then divided into low- and high-risk groups based on their median risk score. Kaplan Meier (K-M) survival curves conducted using the "survival" R package were employed to assess differences in PFI between two groups. Furthermore, a nomogram method was used to construct a clinical prognosis prediction model.

### 3. Determining the optimal PFI threshold

We utilized Pearson’s test to identify genes that demonstrated a linear relationship with PFI (P<0.05). Kaplan Meier (K-M) survival curves, conducted using the “survival” R package, were employed to determine the optimal cut-off value of each PRGs’ expression. The distribution of PFI thresholds were derived by utilizing the linear correlation equation between gene expression and PFI. The optimal PFI threshold was determined, serving as a cutoff to distinguish between the good prognosis and poor prognosis groups.

### 4. DEGs analyses and pathways enrichment analyses

DEGs were selected based on a screening criterion of |log 2 fold change (FC) > 1| and a false discovery rate (FDR)<0.05 using the “GEO2R” tool (https://www.ncbi.nlm.nih.gov/geo/geo2r/)(20). KEGG pathways enrichment analyses were performed using the “clusterProfiler” R package(21).

### 5. Prediction of drug sensitivity

To predict potential responses to common chemotherapy drugs in each group, we calculated the half-maximal inhibitory concentration (IC50) of conventional chemotherapeutic compounds and targeted drugs in HGSOC patients using the “oncoPredict” R package(22). The compounds were obtained from the Genomics of Drug Sensitivity in Cancer (GDSC2) database(23) which contains 969 cancer cell lines and 297 anti-cancer compounds. Differences in IC50 values between different subsets were evaluated using Wilcoxon signed-rank test, with results showing *p* < 0.05 considered statistically significant.

## Results

### 1. Filtrating the PFI-related predictive signature in HGSOC

The study design was depicted in Figure 1A. In this study, univariate COX regression analysis was performed with PFI as the outcome event. A total of 2870 PRGs were identified. LASSO regression analysis ultimately revealed 10 PRGs (ENSG00000127824.9, ENSG00000232325.3, ENSG00000268080.1, ENSG00000169427.2, ENSG00000230567.3, ENSG00000175315.2, ENSG00000184445.7, ENSG00000261122.2, ENSG00000267469.1, ENSG00000101198.10) (Figure 1).

**Figure 1.**
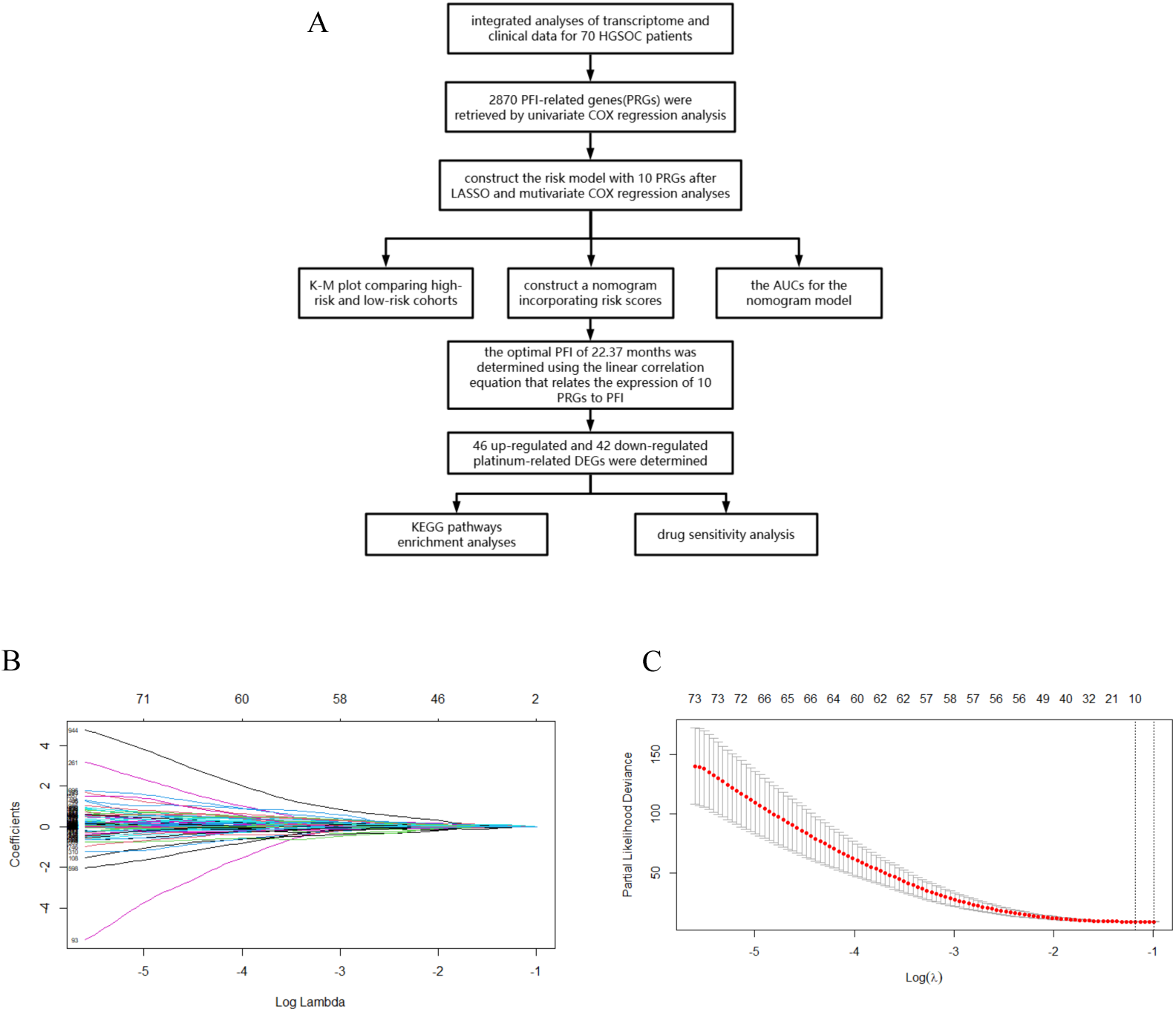
(A) Flow diagram of the study design. (B) LASSO regression analysis was performed with 2870 PFI-associated genes (PRGs). (C) 10 genes were screened to be related to PFI when lambda(**λ**) was min.

### 2. Construction of the PFI-related predictive signature in HGSOC

These ten PRGs include one protective factor (KNTC1) and nine risk factors. Based on transcriptome expression information from the GEO database, the expression levels of these ten PRGs in HGSOC samples were examined. The risk score for HGSOC patients was determined based on their expression levels and regression coefficients: risk score = (0.002158 × TUBA4A expression)+(0.035837 × ENSG00000232325.3 expression)+(0.013061 × ENSG00000268080.1 expression)+(0.017729 × KCNK9 expression)+(0.00752 × ENSG00000230567.3 expression)+(0.069362 × CST6 expression)-(0.00684 × KNTC1 expression)+(0.011352 × LINC02167 expression)+(0.000133 × ENSG00000267469.1 expression)+(0.011322×NKAIN4 expression).

### 3. Assessment of prognostic risk model in HGSOC

Patients were divided into low- and high-risk groups based on the median value of their risk score. Kaplan-Meier analyses revealed that patients in the high-risk cohort had shorter PFI times (Figure 2, *p* < 0.0001). Univariate Cox regression analysis demonstrated that the risk score correlated with patient’s PFI independently while stage and age did not correlate with patient’s PFI. A nomogram was constructed to predict the l-, 3-, and5-year survival rates. The AUCs for the 1- and 3-year nomogram models were 0.903 and 0.901, respectively. Receiver operating characteristic (ROC) curve analysis confirmed that this risk model possesses robust predictive performance independent of other clinicopathological variables (Figure 2).

**Figure 2.**
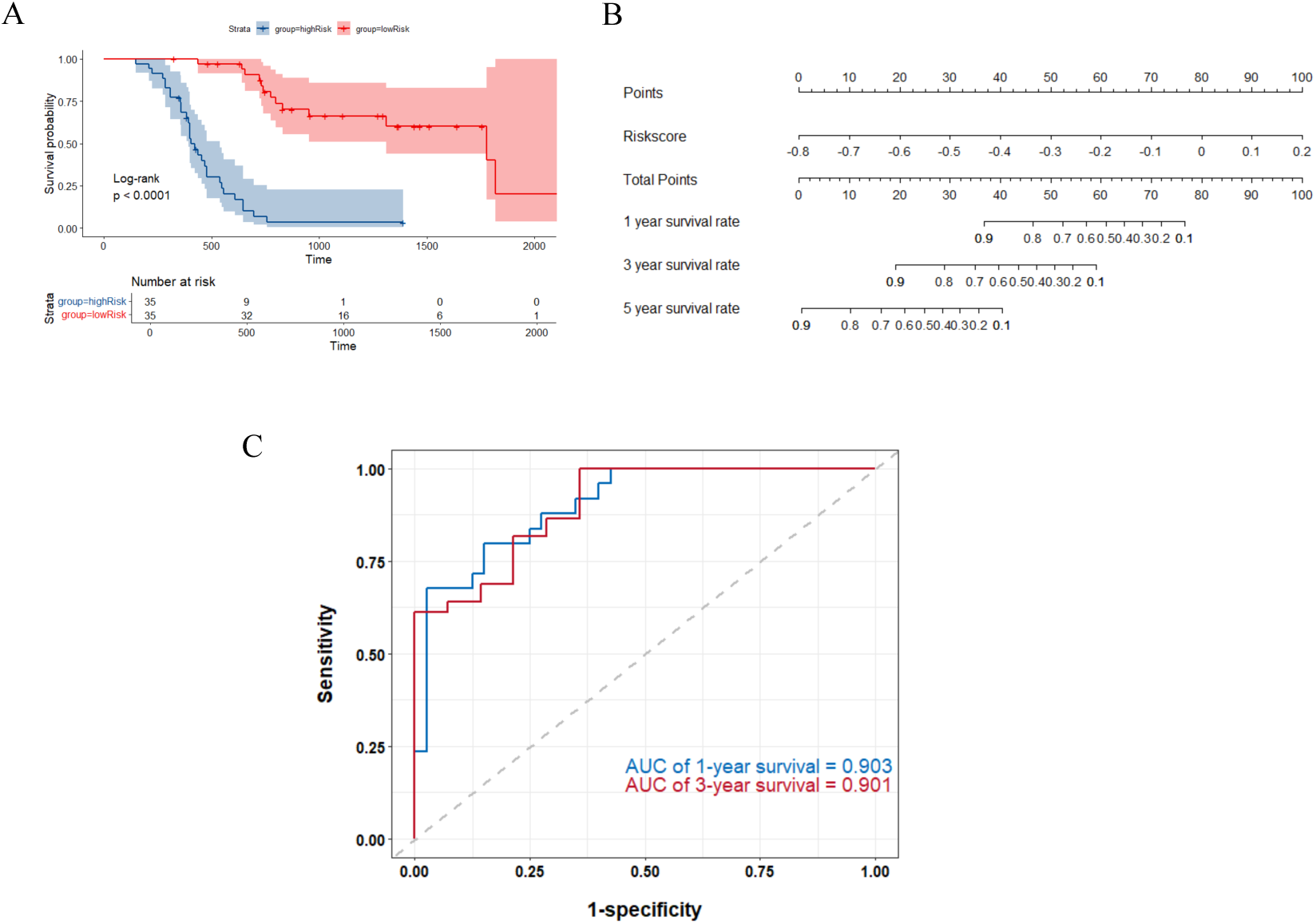
Construction of risk score and prognostic model. (A) Kaplan-Meier curves for the PFI of HGSOC patients in the low- and high-risk groups. (B) A nomogram was constructed based on riskscore identified by multivariate Cox regression analysis, enabling prediction of the recurrence at 1-, 3-, and 5-year after the last platinum-based chemotherapy. (C) The area under the receiver operating characteristic (ROC) curves for the 1- and 3-year intervals were utilized to assess the discriminative ability of the nomogram models.

### 4. The optimal PFI was identified

The Pearson test showed that PRGs’ expression levels exhibited a linear correlation with PFI (p < 0.05) (Figure 3A). K-M survival curves were plotted using each PRG’s expression as a variable (Figure 3B). Cut-off values corresponding to each gene’s PFI data distribution were obtained through its linear relationship with PFI. The overall distribution of PFI was estimated to be 22.37 months (95%CI: 16.97-26.32) (Figure 3C). This time point serves as a cutoff to distinguish between the good prognosis and poor prognosis groups. The PFI distributions for the good prognosis and poor prognosis groups were shown in the volcano plot (Figure 3D). From this result, it can be seen that the PFI within each group was relatively concentrated, meaning that most of the samples in the poor prognosis group have a PFI of less than 22.37 months, while most of the samples in the good prognosis group have a PFI of more than 22.37 months. This further confirms the rationale of our PFI threshold.

**Figure 3.**
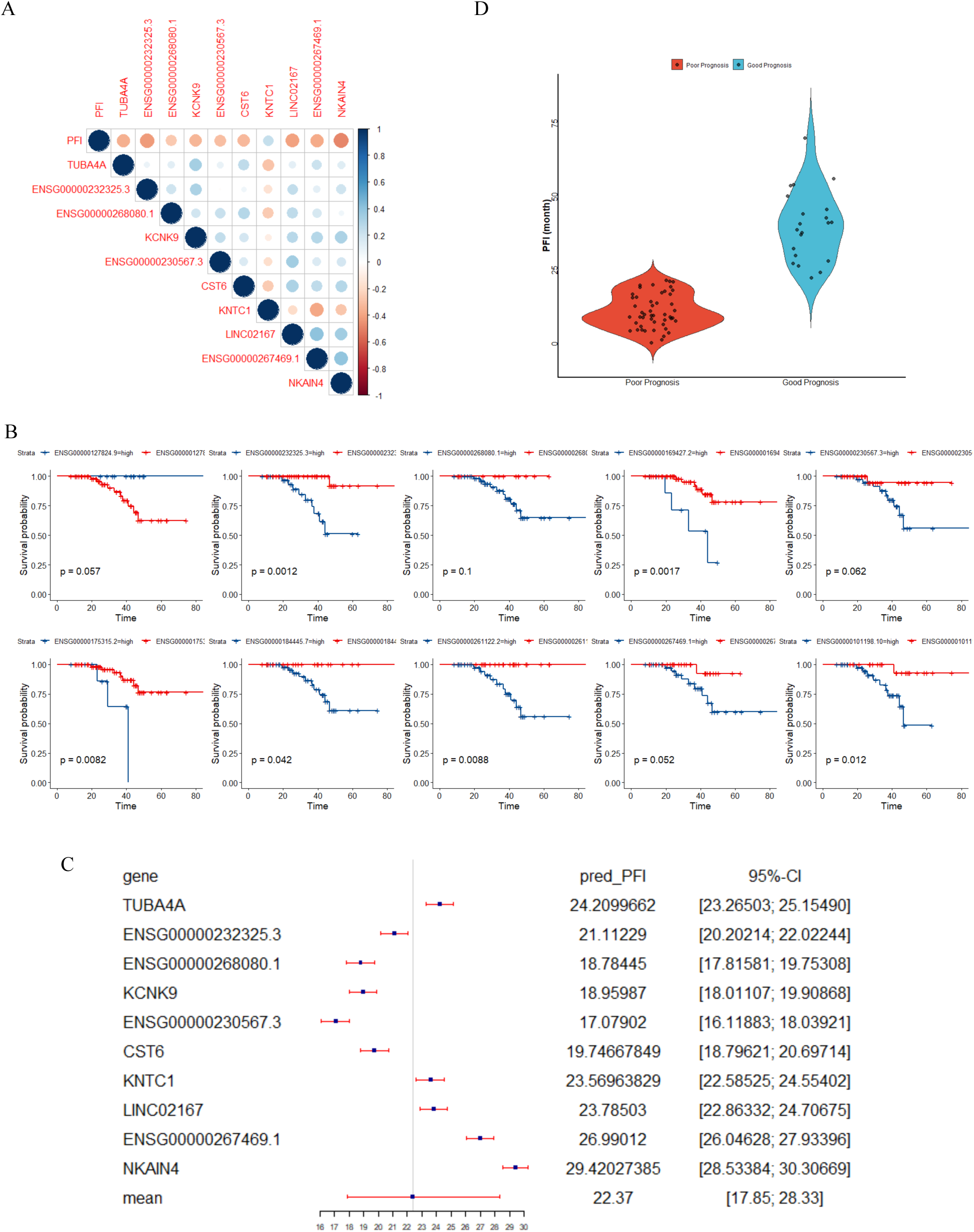
(A) Pearson test were performed between PFI and 10 PRGs. (B) Kaplan-Meier survival curves of PFI for patients in the low- and high-expression groups stratified by each PRG. (C) Cut-off values of PFI-related genes (PRGs)’ expression predicted PFI value by their linear relationship. (D) The distribution of PFI in the high-risk and low-risk groups.

### 5. DEGs analysis

Patients diagnosed with HGSOC were categorized into good prognosis and poor prognosis groups using the PFI threshold (22.37 month we identified above), followed by DEGs analyses. We obtained 88 DEGs in total, consisting of 46 up-regulated DEGs and 42 down-regulated DEGs (p adjust<0.05, |logFC|>1). A volcano plot visualized differences in PRGs’ expressions between groups, and top 10 DEGs were presented in Figure 4.

**Figure 4.**
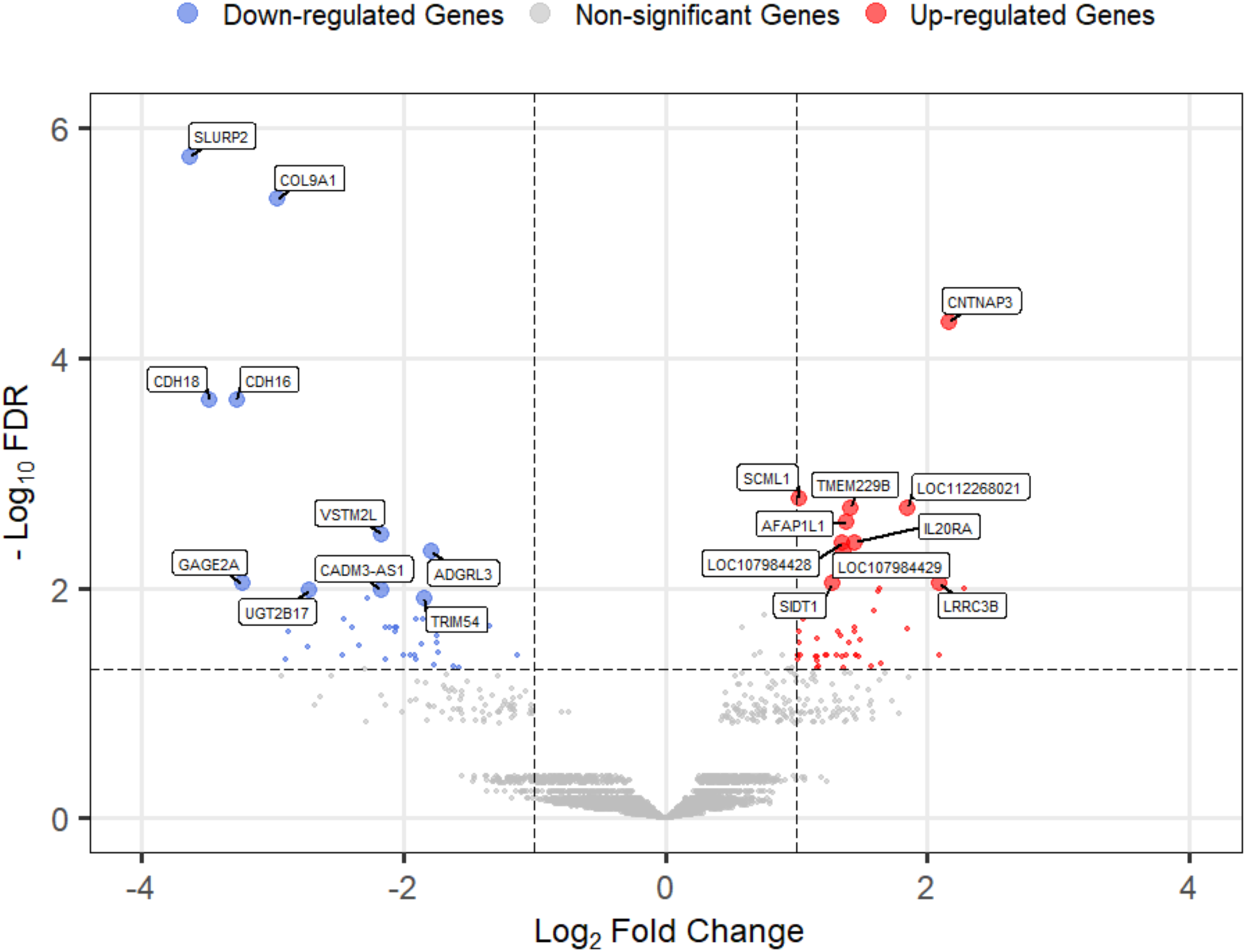
The volcano plot of 88 DEGs. Blue dots represent down-regulated DEGs and red dots represent up-regulated DEGs. Top 10 down-regulated genes, as well as top 10 up-regulated genes were marked.

### 6. KEGG enrichment analysis

KEGG enrichment analyses revealed that the above DEGs were chiefly involved in Melanogenesis, Basal cell carcinoma, Taurine and hypotaurine metabolism, Cushing syndrome, mTOR signaling pathway, Drug metabolism-cytochrome P450 and Bile secretion. The up-regulated DEGs were particularly linked to Taurine and Hypotaurine Metabolism, Melanogenesis, Cushing Syndrome, and mTOR Signaling Pathway. (Figure 5).

**Figure 5.**
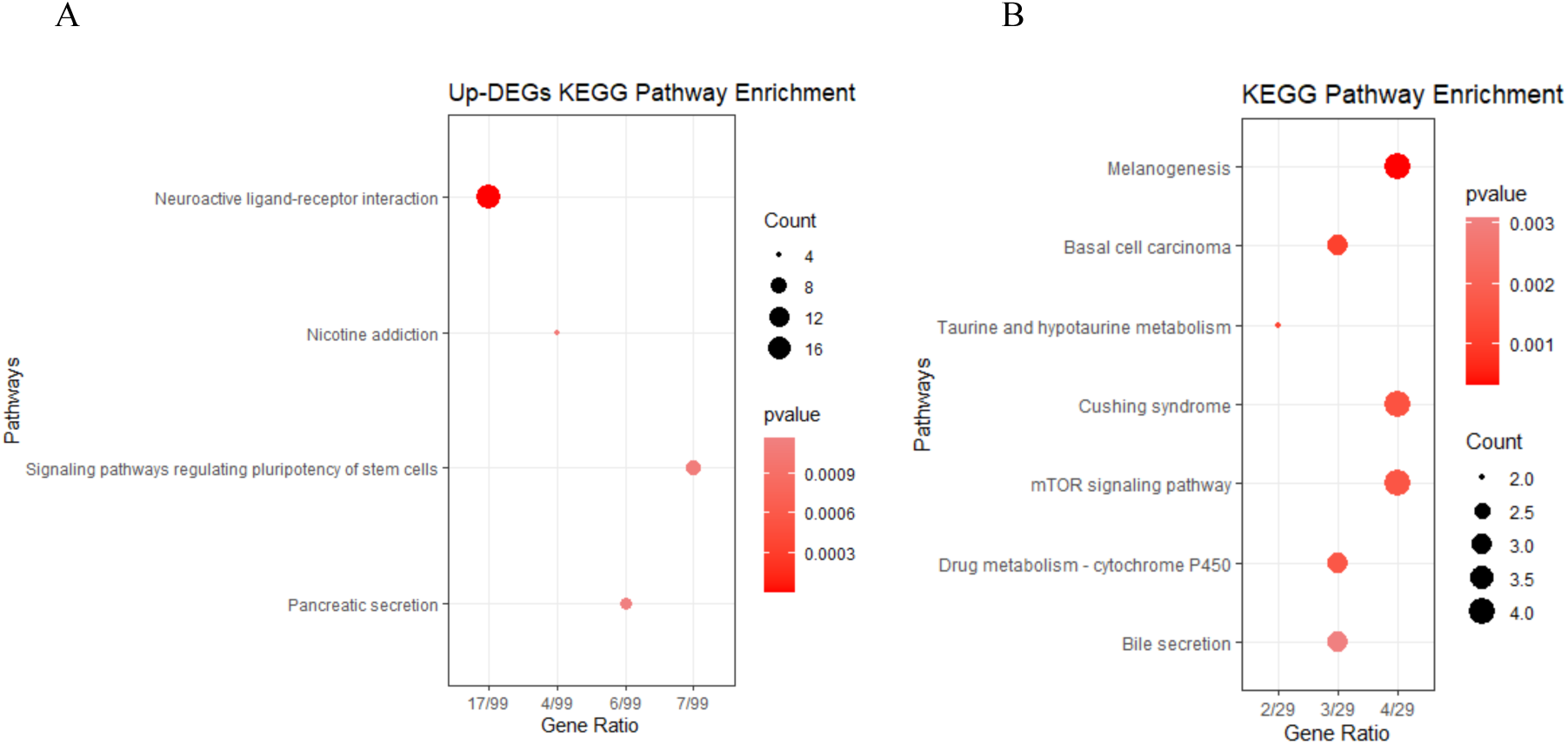
(A) KEGG enrichment analysis of DEGs. (B) KEGG enrichment analysis of up-regulated DEGs.

### 7. Drug sensitivity in HGSOC therapy

We found that the IC50 values of Pevonedistat and GDC0810 were significantly higher in the poor prognosis group than in the good prognosis group. This implies that individuals in the poor prognosis group exhibited reduced susceptibility to these platinum-resistant overcoming medications (Figure 6).

**Figure 6.**
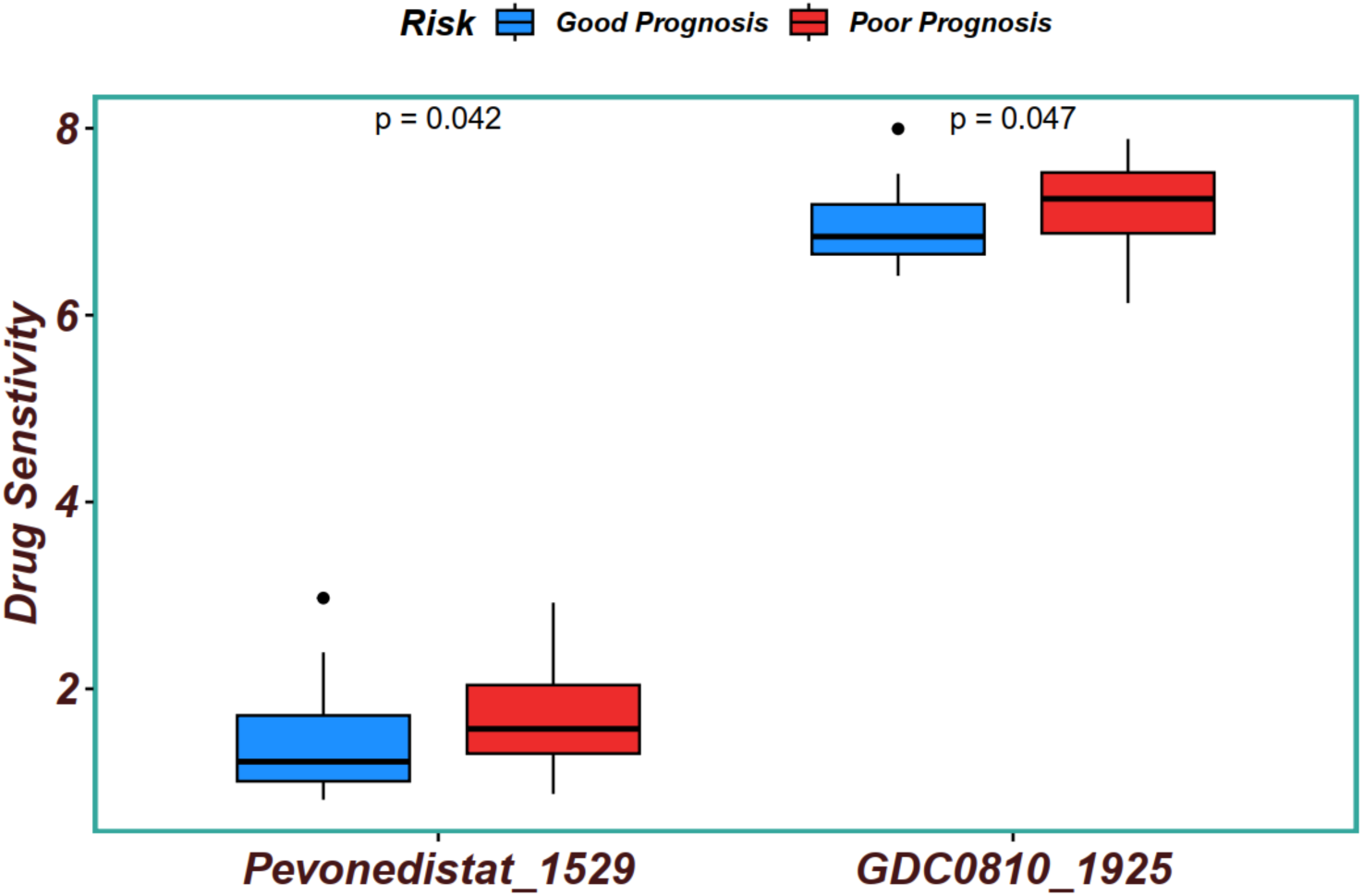
Drug sensitivity analysis. IC50 of Pevonedistat (MLN4924) and GDC0810 in the poor prognosis group and good prognosis group.

## Discussion

Existing evidence suggests that the method of evaluating chemotherapy responses of HGSOC with a PFI of 6 months is increasingly unable to meet clinical requirements. Consequently, our study aimed to identify the optimal PFI threshold, which serves as a cutoff point to differentiate between poor prognosis and good prognosis groups. This could give implications for exploring reliable molecular biomarkers and therapeutic targets for platinum resistance. In our study, the optimal PFI for HGSOC was determined as 22.37 months. With this finding, we further constructed the prognosis prediction model for HGSOC patients and identified ten core PRGs (TUBA4A, ENSG00000232325.3, ENSG00000268080.1, KCNK9, ENSG00000230567.3, CST6, KNTC1, LINC02167, ENSG00000267469.1, NKAIN4). Many of them have been shown to be responsible for platinum resistance. For example, Xu and Xie’s study(24,25) demonstrated that CST6 and KNTC1 can regulate the metastasis and invasion of ovarian cancer. TUBA4A(26) and KCNK9(27) have also been demonstrated to influence the prognosis of solid tumors in lung cancer and breast cancer. These existing findings support the reliability of our analysis results. Notably, the other six genes that we identified here have not yet been studied in platinum-resistance, and most of them are lncRNAs. The roles of lncRNAs in chemoresistance in cancer have been attracted attentions regarding their functions such as in suppression of apoptosis, DNA damage response, epigenetic alterations(28). These genes are thus good candidate targets for platinum resistance which deserve further investigation. Based on the prediction model, we stratified patients into poor prognosis and good prognosis group. The KEGG enrichment analysis revealed that the up-regulated DEGs were particularly enriched in pathways related to taurine and hypotaurine metabolism, melanogenesis, Cushing’s syndrome, and the mTOR signaling pathway. While the involvement of the mTOR signaling pathway in chemoresistance across various cancers is well established(29–31), the roles of the other signaling pathways in chemoresistance in HGSOC merit further investigation.

Moreover, our drug sensitivity analyses indicated that patients in the poor prognosis group are likely sensitive to some chemotherapeutic agents and targeted drugs, such as Pevonedistat (MLN4924) and GDC0810. Neddylation is a posttranslational modification that conjugates a ubiquitin-like protein NEDD8 to substrate proteins, the process of protein neddylation is overactivated in multiple types of human cancers. Pevonedistat selectively inactivates NEDD8 activating enzyme, an E1 enzyme (ubiquitin-activating enzyme), to regulate degradation of intracellular proteins(32). Some investigations identified inhibition of Neddylation as a novel mechanism for overcoming platinum resistance for ovarian cancer in vitro and in vivo(33,34). In addition, there is a Phase I clinical trial of pevonedistat for the treatment of platinum-resistant gastric cancer patients(35). GDC0810 is a molecule inhibitor inactivates ESR1 and ESR2, targeting hormone-related pathways. ESR1 and ESR2 encode two distinct types of estrogen receptors (ERα and ERβ) that play a crucial role in mediating estrogen metabolism and have been identified as genetic factors contributing to the pathogenesis of breast cancer(36). In our recent study, we found that ESR1 plays an important role as a core gene in platinum resistance in HGSOC cancer cells (17). Our findings thus provided implications for personalized chemotherapy treatment for HGSOC patients in the future.

## Conclusions

Our study constructed a prognostic model based on PFI for HGSOC and further explored its implications for chemotherapy resistance. These findings could enhance clinical applications and inform novel anticancer therapeutic strategies targeting HGSOC. We noticed that our study was based on analyses from only one population cohort which could have limitations. Future multi-center analyses with larger sample sizes will provide more universal and comprehensive findings. However, our study offers a new analytical framework for chemotherapy prognosis, which could enhance the ability to classify patients for more effective personalized treatment in clinical practice.

## Data Availability

All data produced in the present work are contained in the manuscript.

https://www.ncbi.nlm.nih.gov/geo/query/acc.cgi?acc=GSE102073

## List of abbreviations

HGSOC: high-grade serous ovarian cancer
PFI: platinum-free interval
PRGs: PFI-related genes
DEGs: differentially expressed genes
GEO: Gene Expression Omnibus
LASSO: least absolute shrinkage and selection operator
K-M: Kaplan Meier
FC: fold change
FDR: false discovery rate
IC50: half-maximal inhibitory concentration
GDSC2: Genomics of Drug Sensitivity in Cancer
ROC: Receiver operating characteristic

## Declarations

### Ethics approval and consent to participate

Not applicable.

### Consent for publication

Not applicable.

### Availability of data and materials

All data generated or analysed during this study are included in this published article. The study used openly available human data that were originally located at: https://www.ncbi.nlm.nih.gov/geo/query/acc.cgi?acc=GSE102073

### Authors’ contributions

LG and GY conceived and supervised the study. YZ and JK handled data curation and analysis, with HX, QL, HT, ZZ, and WH contributing to various aspects of the analysis. SY and GY provided funding support. YZ and LG wrote the manuscript, and all authors approved the final version.

## Acknowledgements

Not applicable.

